# Thromboembolic risk in hospitalised and non-hospitalised Covid-19 patients: A self-controlled case series analysis of a nation-wide cohort

**DOI:** 10.1101/2021.02.02.21251043

**Authors:** Frederick K Ho, Kenneth KS Man, Mark Toshner, Colin Church, Carlos Celis-Morales, Ian CK Wong, Colin Berry, Naveed Sattar, Jill P Pell

**Affiliations:** Institute of Health and Wellbeing, University of Glasgow; School of Pharmacy, University College London; Centre for Safe Medication Practice and Research, Department of Pharmacy and Pharmacology, University of Hong Kong; Department of Medicine, School of Clinical Medicine, University of Cambridge; NHS Greater Glasgow and Clyde; Institute of Cardiovascular & Medical Sciences, University of Glasgow

**Keywords:** Covid-19, thromboembolism, stroke

## Abstract

**Objective:** An unexpectedly large number of people infected with Covid-19 had experienced a thrombotic event. This study aims to assess the associations between Covid-19 infection and thromboembolism including myocardial infarction (MI), ischaemic stroke, deep-vein thrombosis (DVT), and pulmonary embolism (PE).

**Patients and Methods:** A self-controlled case-series study was conducted covering the whole of Scotland’s general population. The study population comprised individuals with confirmed (positive test) Covid-19 and at least one thromboembolic event between March 2018 and October 2020. Their incidence rates during the risk interval (5 days before to 56 days after the positive test) and the control interval (the remaining periods) were compared intra-personally.

**Results:** Across Scotland, 1,449 individuals tested positive for Covid-19 and experienced a thromboembolic event. The risk of thromboembolism was significantly elevated over the whole risk period but highest in the 7 days following the positive test (IRR 12.01, 95% CI 9.91-14.56) in all included individuals. The association was also present in individuals not originally hospitalised for Covid-19 (IRR 4.07, 95% CI 2.83-5.85). Risk of MI, stroke, PE and DVT were all significantly higher in the week following a positive test. The risk of PE and DVT was particularly high and remained significantly elevated even 56 days following the test.

**Conclusion:** Confirmed Covid-19 infection was associated with early elevations in risk with MI, ischaemic stroke, and substantially stronger and prolonged elevations with DVT and PE both in hospital and community settings. Clinicians should consider thromboembolism, especially PE, among people with Covid-19 in the community.

## Introduction

Increasing evidence suggests a potential link between Covid-19 infection and thromboembolism, which could affect a range of organs resulting in: myocardial infarction (MI), ischaemic stroke, pulmonary embolism (PE), and deep vein thrombosis (DVT).

First indications of a potential link came from a case report that described pulmonary embolism in a patient infected with Covid-19 who had no relevant risk factors or past medical history.^2^ Subsequently hospital-based case series supported the hypothesis, including ischaemic stroke in five younger (33-49 years) patients who tested positive for Covid-19.^3^ A recent meta-analysis of 3,487 Covid-19 patients from 30 studies produced a 26% pooled incidence of VTE, but concluded that the existing evidence was low-quality and heterogeneous.^5^ Similar findings were reported by another meta-analysis focused on PE and DVT.^6^ VTE has now been recognised as a relatively common complication of Covid-19 and clinical guidelines recommend the use of pharmacological prophylaxis following risk assessment.^7^ However, clinical trials have provided heterogenous findings, potentially depending on the severity of Covid-19.^8,9^

The current evidence, however, is mainly based on crude incidence from hospitalised case series. Since hospitalised patients are a highly-selected minority of those infected with Covid-19, these studies are unrepresentative and not generalisable to the general population.^10^ It is unknown whether people who are asymptomatic or with mild Covid-19 symptoms (non-hospitalised) were also at a higher risk of thromboembolic events. Even in studies comparing thromboembolic risk between individuals with and without Covid-19^11^, unobserved confounding is still a major concern. To address these limitations, we conducted a self-controlled case series study (SCCS) using a national, general population cohort. This method overcomes bias due to unobserved health conditions. Because SCCS is conducted only amount people with any thromboembolic events, we conducted a supplementary cohort analysis to verify the findings.

## Methods

### Data sources

We undertook individual-level record linkage of five health databases covering the whole of Scotland (5.5 million population) between March 2018 and October 2020: The Community Health Index (CHI) register; Electronic Communication of Surveillance in Scotland (ECOSS); Rapid Preliminary Inpatient Data (RAPID); Scottish Morbidity Record 01 (SMR01), and death certificates.

The CHI register provides sociodemographic information (age, sex, area socioeconomic deprivation). Deprivation is measured using the Scottish Index of Multiple Deprivation (SIMD), derived from seven domains – income, education, health, employment, crime, housing, and access to services – and categorised into general population quintiles. ECOSS collects laboratory data on infectious diseases, including test date and result. RAPID collects real-time data on hospitalisation, including dates of admission and discharge, and type of ward, and SMR01 records diseases using International Classification of Diseases (ICD-10) codes and procedures using Office of Population Censuses and Surveys (OPCS-4) codes. Death certificates provide the date and cause (using ICD-10) of all deaths, whether in hospital or the community. The Community Health Index (CHI), a unique identifier, is used across all databases enabling exact matching. We extracted records covering 1 March 2018 to 5 October 2020 inclusive for all databases except the ECOSS Covid-19 test data which covered 1 March 2020 to 5 October 2020. The Scottish data were accessed through the eDRIS, Public Health Scotland and have been utilised in several previous epidemiological studies.^12,13^ Approval for the study was provided by the Public Benefit and Privacy Panel for Health and Social Care (reference 2021-0064).

In the supplementary cohort analysis, all individuals with a Covid-19 test positive were included as the exposed group. For each exposed individual, 10 age-, sex-, and deprivation-matched individuals who did not have a test positive were included using probability density matching.

### Outcomes

This study included five outcomes ascertained from SMR01 and death certificates: myocardial infarction (MI; ICD-10: I21), ischaemic stroke (I63-64), pulmonary embolism (PE; I26), and deep-vein thrombosis (DVT; I80.1-80.9, I82.8, I82.9), as well as thromboembolism (composite of all four). To test the specificity of any association between Covid-19 and thromboembolism, we also included a composite negative control outcome of elective surgery for hernia repair (OPCS-4 T19, T21-27), colonoscopy (OPCS-4 H22, H25, H28), cataract surgery (OPCS-4 C71-75, C77, C79), or hip/knee replacement (OPCS-4 W37-42, W93-95, O18).

### Statistical Analyses

The self-controlled case series (SCCS) method was chosen to analyse the association between Covid-19 infection and outcomes (Supplementary Figure 1), in favour of a traditional cohort approach, because of its ability to control for intrapersonal time-invariant confounders, and the UK’s testing strategy. Frail individuals with long-term conditions were more likely both to be tested and experience adverse outcomes. These confounders may not be well recorded in the routine data. With a new condition, such as Covid-19, other unknown confounders may also exist. The SCCS method eliminates intrapersonal time-invariant confounders because each person acts as their own control.^14^ The method has been widely-used in epidemiological studies, including influenza and myocardial infarction.^15^

The study population comprised everyone in Scotland who had confirmed (positive real-time PCR test) Covid-19 infection and had experienced one or more thromboembolic event over the study period. The incidence rate ratio (IRR) of thromboembolic outcomes was derived from the ratio of incidence rates in risk and control intervals. The risk interval was defined as between 5 days before and 54 days after the sample was taken for their first positive Covid-19 test. The risk interval was categorised into: 5 to 1 day before; 0 to 7 days after; 8 to 28 days after; and 29 to 56 days after. The five days prior to confirmed infection were included in the risk period to take account of lags in symptom development and testing. The control interval was defined as the remaining study period. Because the UK Covid-19 pandemic started in March 2020, the majority of the control interval occurred prior to infection.

Conditional Poisson regression was used adjusting for participant age in quintile groups, the main time-varying confounder. Deriving rates for both the risk and control intervals from the same individual obviated the need to control statistically for time-invariant confounders. Because individuals who had fatal events prior to the pandemic had not had a chance for Covid-19 test, standard SCCS cannot be applied to fatal events, and the models were run initially for non-fatal hospitalisations. We then repeated the analyses for the composite outcome of hospitalisation or death using the extended SCCS for event-dependent observation periods, which was described elsewhere.^16^

Subgroup analyses were conducted by Covid-19 admission (those with Covid-19 as primary diagnosis versus those without), age (≤75 versus >75 years), sex, and socioeconomic deprivation (SIMD quintile 1-3 versus SIMD quintile 4-5). P-values for subgroup differences were calculated. Additional subgroup analysis was conducted for age (≤65, 66-80, >80 years) to explore any age trends, even though the number of events were not sufficient to conduct formal tests. Three sensitivity analyses were conducted. Firstly, seasonality, in three-month categories, were adjusted because cardiovascular diseases exhibit seasonal patterning. Secondly, we included an extended risk interval, 14 to 6 days prior to a positive test. If the elevated risk in this extended interval is lower than that in the immediate pre-test interval, reverse causation is less likely. Thirdly, as Covid-19 infection was not tested prior to the 2020 pandemic, we restricted the analysis to cases with events after 1 February 2020. Lastly, we calculated the E-values to investigate how robust our findings are regarding time-varying confounders.^17^ A high E-value suggest that only strong time-varying confounder could nullify the findings.

A supplementary cohort analysis was conducted. Time-to-event (from test positive in the exposed individual) to the thromboembolic events was regressed by Covid-19 test positive, controlling for age, sex, and deprivation using Cox proportional hazard model. Proportional hazard assumptions were checked using the Schoenfeld residuals. All analyses were conducted in R version 3.5.1 with the packages *SCCS* and *survival*.

## Results

Of the 30,709 individuals who had at least one positive Covid-19 test (Figure 1) between 1 March 2020 and 5 October 2020, the incidence rates were 44.0, 67.0, 48.6, 18.8 per 1,000 person-years for MI, ischaemic stroke, PE, and DVT respectively. Ths SCCS analysis further excluded 29,260 individuals because they did not have thromboembolic events in the study period. Of the 1,449 individuals who had thromboembolic events, 117 died out-of-hospital, 81 died in-hospital and 1,251 had non-fatal events. Less than one-third (31.5%) of the individuals had a Covid-19 primary diagnosis in hospital. Among people with non-fatal events, the median age were 77 years (interquartile range [IQR] 65-85 years), half were male, and 26.46% lived in the most deprived quintile (Table 1). Median age was older for ischaemic stroke (82 years) and younger for PE (71 years) and DVT (73 years). Women accounted for a higher percentage (58.6%) of those with DVT.

**Figure 1.**
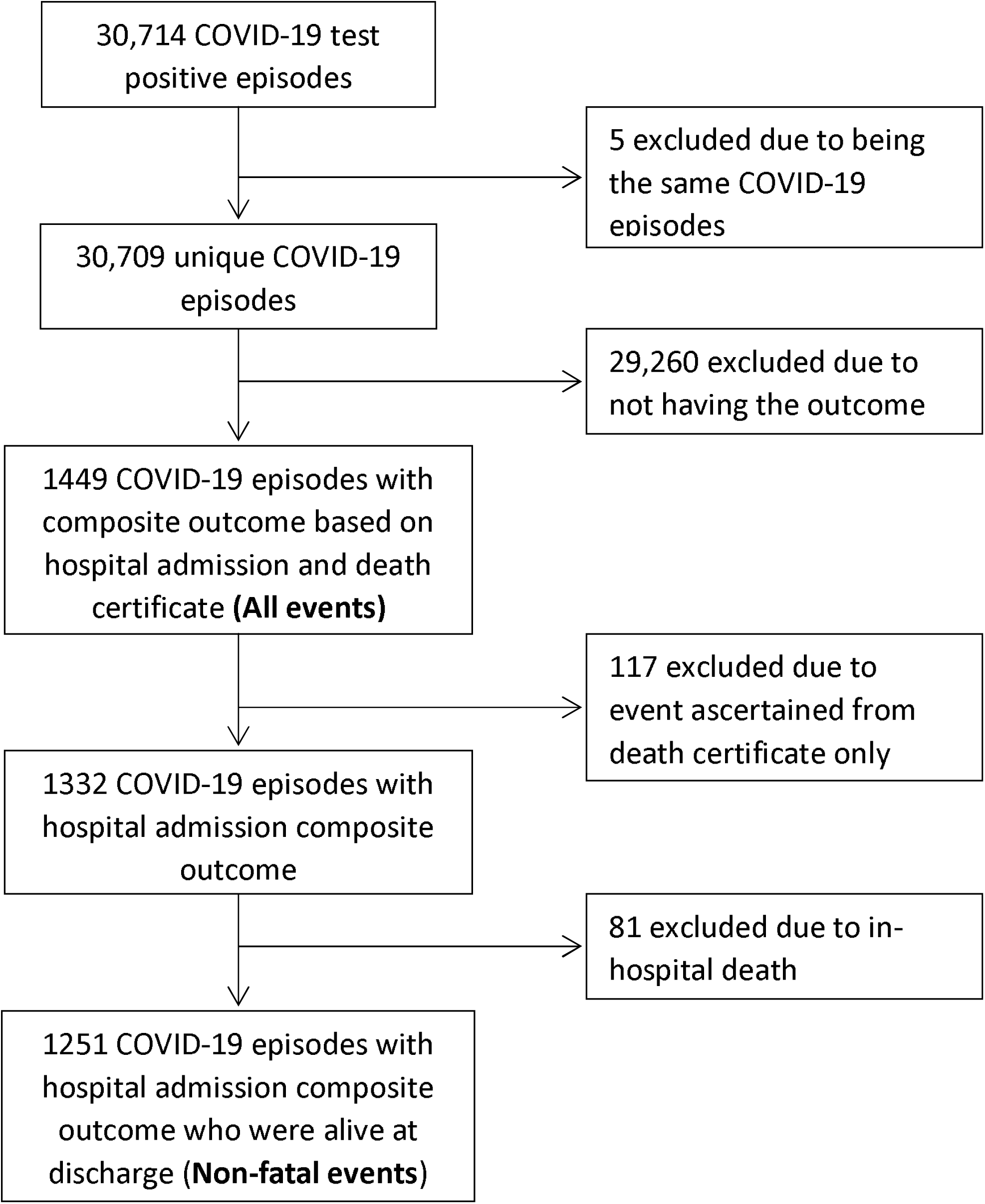
Participant flowchart.

**Table 1.** Patients characteristics for analysis of non-fatal admissions.

|  | **Composite** | **Myocardial infarction** | **Ischaemic stroke** | **Pulmonary embolism** | **Deep-vein thrombosis** | **Elective surgery\*** |
| --- | --- | --- | --- | --- | --- | --- |
| N for all events | 1449 | 376 | 560 | 417 | 179 | 123 |
| N for admissions only | 1332 | 337 | 505 | 391 | 174 | 123 |
| N for non-fatal admissions only | 1251 | 319 | 473 | 359 | 169 | 116 |
| Covid-19 as primary diagnosis in admission episode | 389 (31.5) | 104 (32.6) | 123 (26.0) | 145 (40.4) | 41 (26.6) | 14 (12.1) |
| Median (IQR) age, years | 77 (65-85) | 78 (67-85) | 82 (73-87) | 71 (59-81) | 73 (59-82) | 78 (70-85) |
| Sex |  |  |  |  |  |  |
| Female | 626 (50.04) | 128 (40.13) | 246 (52.01) | 180 (50.14) | 99 (58.58) | 45 (38.79) |
| Male | 625 (49.96) | 191 (59.87) | 227 (47.99) | 179 (49.86) | 70 (41.42) | 71 (61.21) |
| SIMD quintile |  |  |  |  |  |  |
| 1<sup>st</sup> (Most deprived) | 331 (26.46) | 91 (28.53) | 124 (26.22) | 84 (23.40) | 47 (27.81) | 34 (29.31) |
| 2<sup>nd</sup> | 282 (22.54) | 79 (24.76) | 100 (21.14) | 88 (24.51) | 32 (18.93) | 21 (18.10) |
| 3<sup>rd</sup> | 230 (18.39) | 55 (17.24) | 94 (19.87) | 65 (18.11) | 33 (19.53) | 21 (18.10) |
| 4<sup>th</sup> | 230 (18.39) | 53 (16.61) | 95 (20.08) | 68 (18.94) | 27 (15.98) | 25 (21.55) |
| 5<sup>th</sup> (Least deprived) | 178 (14.23) | 41 (12.85) | 60 (12.68) | 54 (15.04) | 30 (17.75) | 15 (12.93) |

Numbers (%) are presented unless otherwise specified.
\*Elective surgery included hernia repair, colonoscopy, cataract surgery, and hip and knee replacement, and is a negative control outcome

The risk of non-fatal thromboembolism was significantly higher over the whole risk interval and highest within the seven days following the positive test (IRR 12.01, 95% CI 9.91-14.56) (Table 2). The associations were strongest for PE followed by DVT (Figure 2); which had similar risk patterns to overall thromboembolism. The associations with MI and ischaemic stroke were smaller in magnitude but nonetheless significant in the 7 days following a positive test, as well as the previous 5 days for MI only. Except for MI, all IRRs in the seven-day post-test interval were significantly stronger than those in the pre-test intervals (Ps <0.04). As expected, there was no significant change in the risk of elective surgery before or after a positive Covid-19 test. The findings for the composite outcome of fatal and non-fatal thromboembolism were similar to those for non-fatal thromboembolism, after accounting for censoring.

**Table 2.** Associations between COVID-19 and outcomes.

| Outcome by risk intervals | Non-fatal events |  | All events<sup>†</sup> |  |
| --- | --- | --- | --- | --- |
|  | IRR (95% CI) | P | IRR (95% CI) | P |
| **Composite** |  |  |  |  |
| 5-1 days before | **4.77 (3.20, 7.10)** | **<0.0001** | **3.71 (2.50, 5.49)** | **<0.0001** |
| 0-7 days after | **12.01 (9.91, 14.56)** | **<0.0001** | **5.70 (4.72, 6.89)** | **<0.0001** |
| 8-28 days after | **2.82 (2.16, 3.67)** | **<0.0001** | **1.54 (1.22, 1.94)** | **0.0003** |
| 28-56 days after | **2.30 (1.77, 3.00)** | **<0.0001** | **1.51 (1.21, 1.88)** | **0.0002** |
| **Myocardial infarction** |  |  |  |  |
| 5-1 days before | **5.15 (2.54, 10.46)** | **<0.0001** | **3.79 (1.86, 7.71)** | **0.0002** |
| 0-7 days after | **5.16 (3.04, 8.73)** | **<0.0001** | **1.98 (1.23, 3.18)** | **0.005** |
| 8-28 days after | 1.51 (0.77, 2.95) | 0.23 | 0.85 (0.50, 1.44) | 0.55 |
| 28-56 days after | 1.15 (0.56, 2.35) | 0.70 | 0.90 (0.53, 1.50) | 0.67 |
| **Ischaemic stroke** |  |  |  |  |
| 5-1 days before | 2.12 (0.88, 5.13) | 0.10 | 1.58 (0.65, 3.84) | 0.31 |
| 0-7 days after | **7.22 (5.02, 10.38)** | **<0.0001** | **3.25 (2.34, 4.50)** | **<0.0001** |
| 8-28 days after | 0.75 (0.35, 1.58) | 0.45 | 0.69 (0.42, 1.12) | 0.14 |
| 28-56 days after | 1.11 (0.63, 1.94) | 0.72 | 0.94 (0.63, 1.41) | 0.77 |
| **Pulmonary embolism** |  |  |  |  |
| 5-1 days before | **9.95 (5.42, 18.27)** | **<0.0001** | **7.47 (4.13, 13.51)** | **<0.0001** |
| 0-7 days after | **27.55 (20.55, 36.95)** | **<0.0001** | **16.81 (12.46, 22.69)** | **<0.0001** |
| 8-28 days after | **7.27 (5.07, 10.43)** | **<0.0001** | **4.52 (3.21, 6.35)** | **<0.0001** |
| 28-56 days after | **5.59 (3.87, 8.07)** | **<0.0001** | **3.54 (2.54, 4.93)** | **<0.0001** |
| **Deep vein thrombosis** |  |  |  |  |
| 5-1 days before | **4.67 (1.48, 14.72)** | **0.008** | **4.23 (1.34, 13.32)** | **0.01** |
| 0-7 days after | **17.44 (11.00, 27.66)** | **<0.0001** | **11.51 (7.30, 18.16)** | **<0.0001** |
| 8-28 days after | **3.64 (1.90, 7.01)** | **0.0001** | **2.43 (1.27, 4.67)** | **0.008** |
| 28-56 days after | 1.98 (0.91, 4.29) | 0.08 | 1.77 (0.92, 3.42) | 0.09 |
| **Elective surgeries\*** |  |  |  |  |
| 5-1 days before | - | - | - | - |
| 0-7 days after | 1.69 (0.41, 6.88) | 0.47 | 1.28 (0.40, 4.06) | 0.67 |
| 8-28 days after | 1.78 (0.65, 4.90) | 0.26 | 0.94 (0.34, 2.59) | 0.91 |
| 28-56 days after | 2.28 (0.98, 5.32) | 0.06 | 1.19 (0.51, 2.76) | 0.68 |

Patients' age quintile was adjusted
IRR: incidence rate ratio
\*Elective surgery included hernia repair, colonoscopy, cataract surgery, and hip/knee replacement, and is a negative control outcome
<sup>†</sup>Including both fatal and non-fatal events, with event dependent observation handled using specialised method

**Figure 2.**
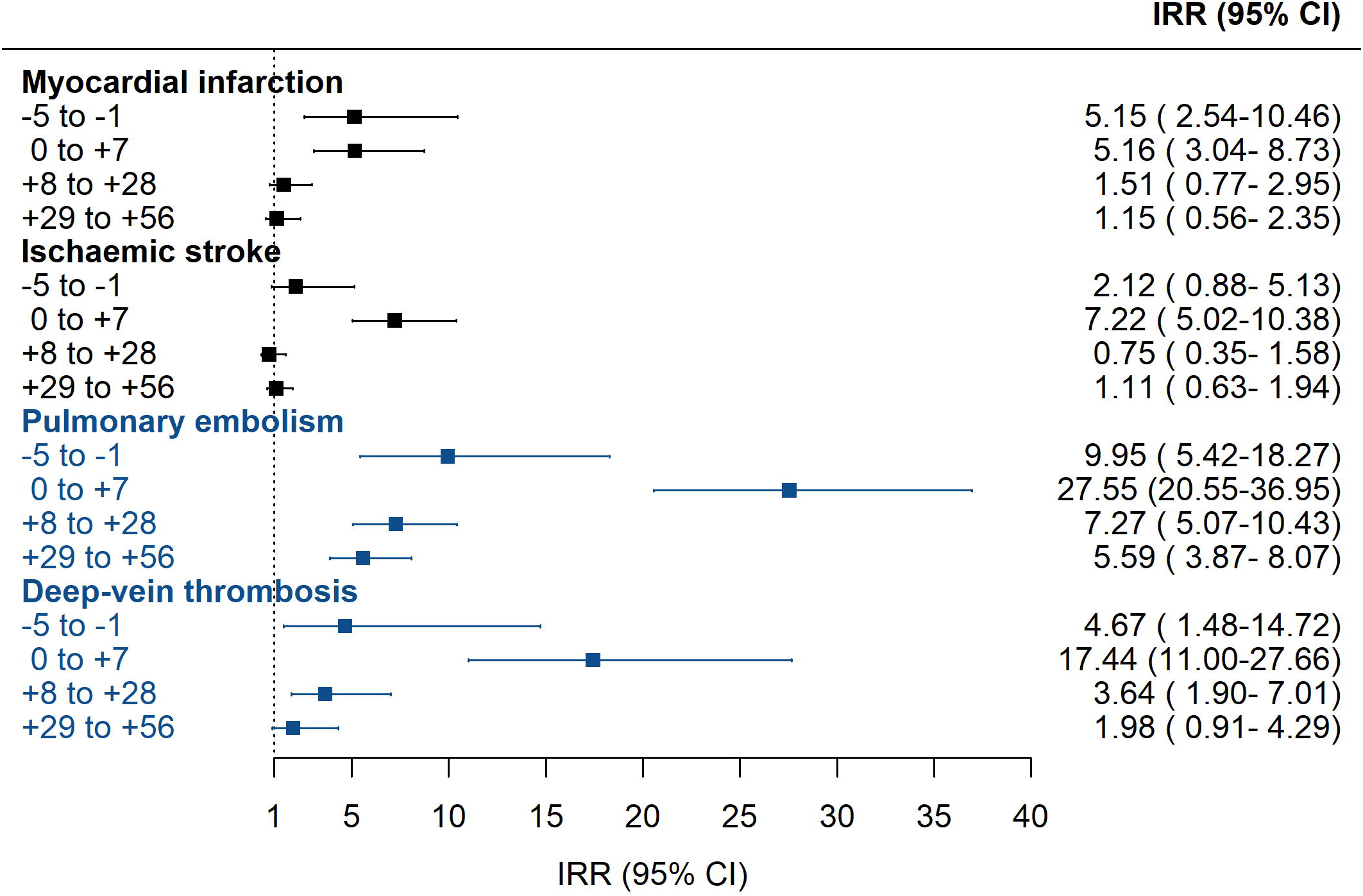
Associations between Covid-19 and non-fatal outcomes. IRR shown is the within incidence rate ratio for outcomes. Incidence rates in the risk period (5 days prior to 56 after a positive Covid-19 test) were compared against the control period (all remaining time in study period) for each person.

Adjusting for seasonality did not alter the findings (Supplementary Table 1). The extended pre-test risk interval generally had lower IRRs than the immediate pre-test interval, and were non-significant for MI, ischaemic stroke, and PE. Including only participants with thromboembolic events after February 2020 resulted in similar IRR estimates. The E-values ranged from 5.53 (MI) to 40.59 (PE) for the lower bound of 95% CIs within seven days of a positive test (Supplementary Table 2).

On subgroup analysis, the associations between a test positive and thromboembolism were significant regardless of Covid-19 admission, even though the elevation of risk was stronger among those admitted for Covid-19 (Table 3). A positive Covid-19 test was also associated with higher risk of thromboembolism regardless of age, but the magnitude of risk was significantly higher (P_interaction_ <0.0001) in people younger than 75 years. Compared with people aged older than 75 years, those younger had 23 and 47 times higher elevated thromboembolism and PE risk, respectively, within seven days of a positive Covid-19 test (Table 3). There appears to be a dose-response trend by age even though insufficient sample size inhibited formal testing (Supplementary Table 2). A positive Covid-19 test was associated with higher risk of overall thromboembolism, PE and DVT in both women and men, but the magnitude of risk was higher in men (P_interaction_ <0.006). The association between a positive Covid-19 test and ischaemic stroke was significant in men only. There was no consistent evidence of socioeconomic deprivation being an effect modifier (Supplementary Table 3).

**Table 3.** Subgroup analysis for non-fatal events.

| Outcome by risk intervals | Covid-19 hospitalisation |  |  | Age |  |  | Sex |  |  |
| --- | --- | --- | --- | --- | --- | --- | --- | --- | --- |
|  | Yes<br>IRR (95% CI) | No<br>IRR (95% CI) | P<sub>interaction</sub> | ≤75 years<br>IRR (95% CI) | >75 years<br>IRR (95% CI) | P<sub>interaction</sub> | Female<br>IRR (95% CI) | Male<br>IRR (95% CI) | P<sub>interaction</sub> |
| **Composite** |  |  |  |  |  |  |  |  |  |
| 5-1 days before | **12.45 (7.37, 21.03)** | **2.48 (1.33, 4.63)** | **0.0001** | 3.80 (2.44, 5.93) | 2.60 (1.81, 3.72) | 0.19 | **2.22 (1.41, 3.47)** | **4.31 (3.02, 6.15)** | **0.03** |
| 0-7 days after | **36.97 (28.69, 47.64)** | **4.07 (2.83, 5.85)** | **<0.0001** | **22.78 (17.58, 29.53)** | **5.94 (4.35, 8.12)** | **<0.0001** | **6.36 (4.47, 9.04)** | **19.44 (15.38, 24.58)** | **<0.0001** |
| 8-28 days after | **6.16 (4.18, 9.09)** | **1.82 (1.26, 2.63)** | **<0.0001** | **5.79 (4.16, 8.07)** | **1.24 (0.78, 1.97)** | **<0.0001** | 2.64 (1.83, 3.82) | 3.19 (2.18, 4.66) | 0.50 |
| 28-56 days after | **4.85 (3.27, 7.20)** | **1.50 (1.04, 2.17)** | **<0.0001** | **4.27 (3.03, 6.03)** | **1.12 (0.72, 1.74)** | **<0.0001** | 2.28 (1.59, 3.27) | 2.46 (1.66, 3.65) | 0.79 |
| **Myocardial infarction** |  |  |  |  |  |  |  |  |  |
| 5-1 days before | 3.58 (0.86, 14.88) | 6.17 (2.73, 13.95) | 0.52 | 4.14 (2.00, 8.54) | 3.36 (1.80, 6.26) | 0.67 | 4.29 (2.15, 8.58) | 3.47 (1.82, 6.60) | 0.66 |
| 0-7 days after | 8.09 (3.87, 16.90) | 3.38 (1.50, 7.65) | 0.12 | 6.19 (2.85, 13.42) | 3.65 (1.69, 7.88) | 0.34 | 3.35 (1.22, 9.18) | 6.45 (3.46, 12.00) | 0.28 |
| 8-28 days after | 1.00 (0.24, 4.17) | 1.85 (0.86, 3.96) | 0.46 | 2.49 (1.08, 5.77) | 0.77 (0.24, 2.45) | 0.11 | 1.58 (0.57, 4.39) | 1.43 (0.58, 3.50) | 0.89 |
| 28-56 days after | 1.28 (0.39, 4.22) | 1.15 (0.47, 2.81) | 0.89 | 1.01 (0.32, 3.23) | 0.97 (0.35, 2.64) | 0.96 | 1.41 (0.51, 3.92) | 0.94 (0.34, 2.57) | 0.60 |
| **Ischaemic stroke** |  |  |  |  |  |  |  |  |  |
| 5-1 days before | 3.23 (0.79, 13.30) | 1.62 (0.52, 5.06) | 0.45 | 1.70 (0.54, 5.39) | 1.84 (1.00, 3.37) | 1.00 | 1.21 (0.50, 2.96) | 2.58 (1.32, 5.08) | 0.18 |
| 0-7 days after | **14.03 (8.11, 24.27)** | **4.22 (2.50, 7.12)** | **0.0019** | **17.81 (10.67, 29.72)** | **3.63 (2.07, 6.38)** | **<0.0001** | **2.05 (0.84, 5.00)** | **13.27 (8.79, 20.04)** | **0.0002** |
| 8-28 days after | 1.26 (0.39, 4.07) | 0.51 (0.19, 1.37) | 0.25 | 0.45 (0.06, 3.27) | 0.81 (0.36, 1.84) | 1.00 | 0.75 (0.28, 2.03) | 0.72 (0.23, 2.26) | 0.96 |
| 28-56 days after | 2.37 (1.06, 5.29) | 0.60 (0.27, 1.36) | 0.02 | 1.46 (0.53, 4.02) | 0.93 (0.47, 1.83) | 0.47 | 1.20 (0.59, 2.45) | 0.96 (0.39, 2.37) | 0.75 |
| **Pulmonary embolism** |  |  |  |  |  |  |  |  |  |
| 5-1 days before | **50.25 (24.26, 104.07)** | **2.09 (0.52, 8.44)** | **0.0001** | 5.36 (2.60, 11.08) | 3.40 (1.64, 7.03) | 0.39 | **1.98 (0.73, 5.37)** | **8.10 (4.42, 14.86)** | **0.02** |
| 0-7 days after | **135.97 (88.89, 207.98)** | **3.92 (1.83, 8.40)** | **<0.0001** | **46.84 (32.21, 68.12)** | **10.36 (5.99, 17.91)** | **<0.0001** | **15.22 (9.30, 24.90)** | **43.82 (29.78, 64.48)** | **0.001** |
| 8-28 days after | **23.97 (14.03, 40.97)** | **3.54 (2.03, 6.16)** | **<0.0001** | **12.64 (8.20, 19.49)** | **2.04 (0.93, 4.48)** | **0.0001** | 5.77 (3.42, 9.71) | 9.35 (5.63, 15.54) | 0.20 |
| 28-56 days after | **16.26 (9.35, 28.27)** | **2.99 (1.74, 5.15)** | **<0.0001** | **8.13 (5.16, 12.81)** | **1.88 (0.90, 3.94)** | **0.0009** | 4.98 (3.02, 8.21) | 6.21 (3.58, 10.77) | 0.57 |
| **Deep vein thrombosis** |  |  |  |  |  |  |  |  |  |
| 5-1 days before | **24.07 (5.54, 104.49)** | **1.82 (0.25, 13.20)** | **0.04** | 2.20 (0.54, 9.04) | 3.83 (1.51, 9.69) | 1.00 | 3.18 (1.16, 8.73) | 3.90 (1.20, 12.65) | 0.80 |
| 0-7 days after | **92.44 (46.02, 185.68)** | **3.77 (1.36, 10.46)** | **<0.0001** | 24.21 (13.13, 44.64) | 10.59 (5.11, 21.97) | 0.09 | **8.04 (3.49, 18.57)** | **33.91 (18.80, 61.18)** | **0.006** |
| 8-28 days after | 9.54 (3.15, 28.93) | 2.75 (1.18, 6.40) | 0.08 | 5.30 (2.38, 11.80) | 1.74 (0.53, 5.68) | 0.13 | 4.89 (2.33, 10.26) | 2.03 (0.49, 8.45) | 0.33 |
| 28-56 days after | 1.46 (0.19, 11.55) | 2.35 (1.01, 5.46) | 0.67 | 2.85 (1.11, 7.28) | 0.91 (0.22, 3.82) | 0.19 | 1.41 (0.44, 4.50) | 3.49 (1.23, 9.90) | 0.26 |

Patients' age quintile was adjusted
IRR: incidence rate ratio; SIMD: Scottish Index of Multiple Deprivation

The findings from cohort analysis were consistent with those from SCCS (Supplementary Table 4). Individuals who had a Covid-19 infection was at a higher risk of all of the outcomes, with strongest association with PE (HR 24.04, 95% CI 18.49-31.33), followed by DVT (HR 10.45, 95% CI 7.02-15.56), ischaemic stroke (HR 4.40, 95% CI 3.44-5.63), and MI (HR 3.31, 95% CI 2.59-4.22).

## Discussion

In this national, general population study including hospitalised and community-dwelling individuals, we demonstrated an elevated risk of thromboembolism in temporal proximity to confirmed Covid-19 infection. In the week following a positive test, participants were at significantly increased risk of MI, ischaemic stroke, PE and DVT, with the increased risk of the latter two being marked (Day 0 to +7 IRRs of >27 and >17-fold, respectively) – with risk ratios substantially exceeding those previously associated with upper respiratory infections^18^ – and elevated risk continuing for some time thereafter. The risk ratios were even higher in younger people and in men. The clear implication of this work is that PE/DVT risks are substantially elevated in hospitalised patients as compared to more modest and shorter atherothrombotic risks. However, there appears a broader thrombotic impact not confined to hospitalised populations, albeit at a lower risk level.

It is worth noting that the associations were also significant in individuals not hospitalised for Covid-19. Although the IRRs were modest compared with the hospitalised group, the excess risk for PE was sustained at near three-fold for more than 1-2 months after the initial Covid-19 infection. This modest excess risk may also be applicable to a large number of people who were infected with Covid-19 but not hospitalised, which could mean a sizeable population burden. The annual incidence of PE in the UK general population was 0.98 per 1,000^19^. If the IRR on this study (3.92 in the first 7 days of non-hospitalised group) is applicable to the general population, this would translate to a rate difference of 3.84 in 1,000. There were 4.27 million people who tested positive for Covid-19 in the UK as of 16 March 2021, indicating that at least 16,400 new PE cases could have been caused by Covid-19.

At the present time, unpublished results from ICU Covid-19 populations have led to early stopping of anticoagulant therapeutic arms because of signals suggestive of harm.^8^ Conversely the same collated international studies have intimated a significant decreased need for life support and improved results from less severe hospitalised patients.^9^ Such heterogenous results could be related to the severity of Covid-19, as well as the timing of administering pharmacologic prophylaxis. Given the potentially treatable nature of thrombotic events, urgent work needs to be considered in prevention and treatment trial design to consider risk stratification strategy that includes Covid-19 severity, age, and sex.

Our new findings are in line but meaningfully extend previous Covid-19 studies, including another national cohort from Danmark^11^. A meta-analysis of over 100,000 Covid-19 patients reported that 1.2% developed ischaemic stroke;^20^ a large proportion even considering their age and vascular risk profile. A hospital-based case-control study of 123 patients found an association (odds ratio 3.9) between Covid-19 infection and acute ischemic stroke, after controlling for age, sex, and vascular risk factors.^21^ Similarly, two meta-analyses reported high rates of PE and DVT in patients with Covid-19.^5,6^ Of note, traditional thromboembolic risk factors were not significantly associated with PE in Covid-19 patients suggesting the pathways may be different.^22^ It should also be noted that previous studies^23^ have shown that the PE found in severe Covid-19 patients might actually be primarily caused by pulmonary thrombi rather than pulmonary emboli, which warrants further investigation.

This study’s association pattern for MI is similar to that for influenza, with 5-6 times higher risk in the first 7 days after a test positive.^15^ However, the association of Covid-19 with VTE appeared to be much stronger than that of other infections. For example, a study using the same SCCS method found the elevated risk of DVT was much lower (IRR 1.91 in the first 2 weeks) for upper respiratory infections.^18^ The same study also found that the risk of PE elevated (IRR 2.11 in the first 4 weeks) following urinary tract infection. These suggest that Covid-19 may have either different mechanisms, or a stronger systemic inflammation (in keeping with the cytokine storm), leading to an exponential difference in the risk of PE/DVT compared to other infections, while having similar elevation in MI risk.

Our study demonstrated that the association with ischaemic stroke was significantly stronger in younger (≤75 years) individuals. This is consistent with previous reports of relatively young people (mean age 53-60 years) with Covid-19 requiring thrombectomy.^24-26^ In addition, among stroke patients, those who tested positive for Covid-19 were on average 7-15 years younger than those tested negative.^27,28^ The underlying mechanism warrants further investigation but could relate to cytokine storm, at least in some people.^29^ Historical reports showed healthy young people were more likely to experience cytokine storm following viral infections,^29^ and cytokine storm in Covid-19 patients leading to hypercoagulable was a hypothesised mechanism for thromboembolism.^30^ The finding that Covid-19 is associated with a higher risk of thromboembolism in men than women may partially explain our previous finding that men have worse case-fatality following Covid-19 infection.^31^ This hypothesis requires further study.

Our study has several strengths. Firstly, it was unselective; covering the whole of Scotland and all confirmed Covid-19 cases regardless of whether they were hospitalised. This avoided the selection bias intrinsic to hospital-based studies. Since both Covid-19 infection and thromboembolism increase the chance of hospitalisation, selecting only hospital cases inevitably results in collider bias.^10^ Secondly, time-invariant confounders, including unknown and unmeasured confounders, were perfectly controlled by using participants as their own controls., The key time-varying confounders, age and seasonality, were adjusted for in the model.^14^ The use of E-values showed that the elevated risk within seven days of test positive would only be meaningfully nullified if there were very strong time-varying confounders that could increase/decrease the risk of test positive and thromboembolic events by 5 to 20 times. Thirdly, we were able to separately analyse non-fatal events, using the standard SCCS method, and all events, using a specific method designed for censored data,^16^ and the two approaches produced consistent findings. This, along with the sensitivity analysis including only events shortly before the Covid-19 pandemic, suggest the results should be robust against immortal time biases.

However, the findings of this study are still subject to the following limitations. To ensure internal validity, this study opted for the SCCS method, which only included patients with at least one thromboembolism during the study period. This may limit the generalisability of the findings to people with lower risk of these events even though our confirmatory cohort analysis showed similar results. It should be noted that, if the elevated risk of PE is truly causal, the estimates that we provided could be an underestimate. The IRR for the latest categories in the risk interval was still significantly greater than one, suggesting a long tail of risk elevation and thus some of the pre- and post-infection control interval could be misspecified. Patients with no or mild symptoms from Covid-19 infection are less likely to have been tested, especially at the beginning of the pandemic when testing capacity was lower. The increased risk of thromboembolism demonstrated in the days prior to confirmed infection is likely to reflect the time lag between actual date of infection and our proxy measure of it; date of specimen collection. Reverse causation is possible in some patients; for example, nosocomial infection of patients hospitalised for thromboembolic events. However, the lack of an association with elective surgery suggests that any reverse causation is unlikely to fully explain our findings. The lowered risk in extended pre-test interval for outcomes except MI also does not support strong reverse causation. It is highly likely that there was underreporting of events from the first wave. There were 1465 individuals who died of suspected Covid-19 (ICD-10: U07.2) without any tests suggesting individuals who had Covid-19 but untested is only a small proportion (4.8%) compared to those tested and unlikely to change our conclusion. Even though there was no role for routine CT scanning in Covid-19^32^ and data on rates of advanced imaging are not yet clear, it is our expectation that more extensive imaging in subsequent waves is highly likely to increase pick-up of thrombus.

In conclusion, Covid-19 infection was associated with substantially elevated risk of PE and DVT, with excess PE risk lasting at least 8 weeks post-infection. These complications should be addressed through prophylaxis and early detection; clinicians should be alerted to the possibility of PEs in community treated patients with residual or prolonged symptoms. Clinical trials to prevent thrombotic events should consider the post-hospital convalescent stage where we have demonstrated ongoing increased risk in addition to younger individuals with Covid-19.

## Data Availability

Relevant data can be requested via eDRIS, Public Health Scotland (https://www.isdscotland.org/products-and-services/edris/).

## Acknowledgements

This study was supported by the Wellcome Trust ISSF COVID Response Fund in the University of Glasgow. The authors would like to acknowledge the support of the eDRIS Team (Public Health Scotland), especially Ms Johanna Bruce, for their involvement in obtaining approvals, provisioning and linking data and the use of the secure analytical platform within the National Safe Haven.

