## Supplementary materials for "Thromboembolic risk in hospitalised and non-hospitalised Covid-19 patients: A self-controlled case series analysis of a nation-wide cohort"

**Supplementary Appendix**

### **Supplementary Figure 1**. Illustrative diagram of the self-controlled case series analysis

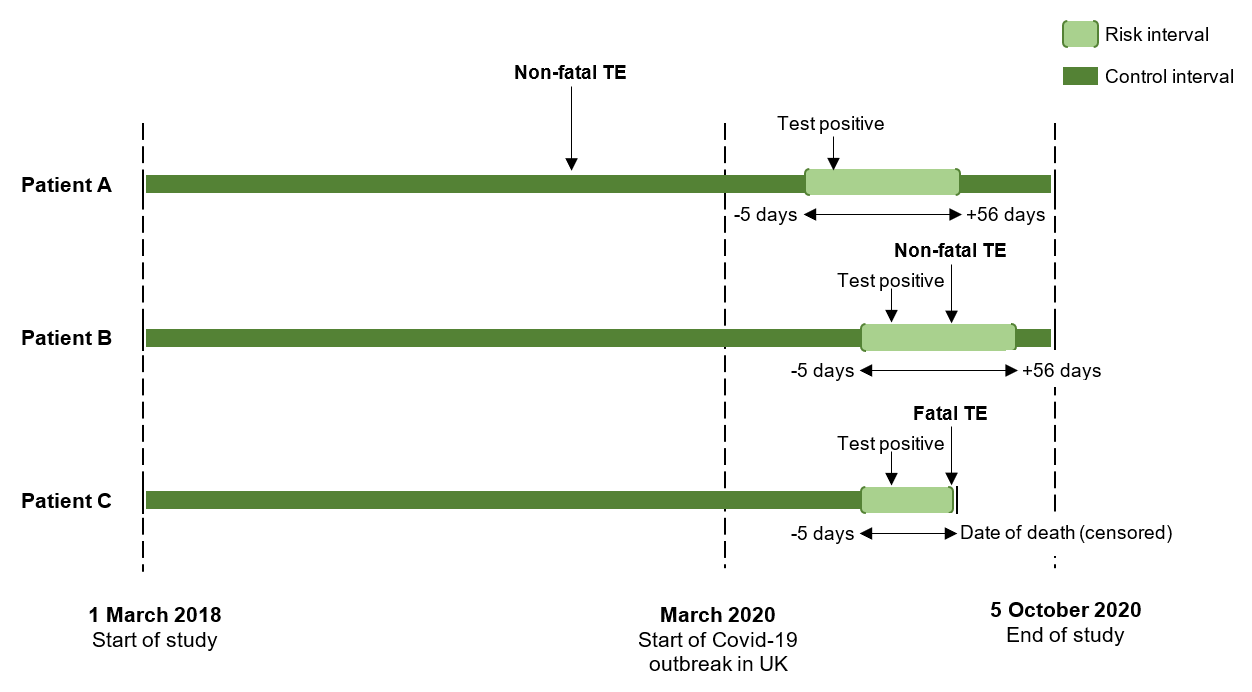

TE: thromboembolism

Self-controlled case series analysis compares the incidence rate of TE during the risk interval (defined as -5 to +56 days of Covid-19 test positive) and that during the control interval (any other study period). Patients A and B represent persons who had non-fatal TE in the risk and control intervals respectively. Patient C represent a person who had fatal TE during the risk interval. This censors the observation, which violates the assumption of ordinary SCCS analysis. Specific method was used to handle analysis including both fatal and non-fatal TEs.

### **Supplementary Table 1**. Sensitivity analyses for the associations between Covid-19 and non-fatal outcomes

| **Outcome by** | **Adjusted for seasonality** | | **Extended pre-test risk interval** | | **Restricted study period (Feb-Oct 2020)** | |
| --- | --- | --- | --- | --- | --- | --- |
| **risk intervals** | **IRR (95% CI)** | **P** | **IRR (95% CI)** | **P** | **IRR (95% CI)** | **P** |
| **Composite** |  |  |  |  |  |  |
| 14-6 days before* | - | - | **2.44 (1.64, 3.63)** | **<0.0001** | - | - |
| 5-1 days before | **3.97 (2.66, 5.94)** | **<0.0001** | **4.36 (2.99, 6.36)** | **<0.0001** | **3.82 (2.52, 5.79)** | **<0.0001** |
| 0-7 days after | **9.81 (7.98, 12.06)** | **<0.0001** | **12.26 (10.11, 14.87)** | **<0.0001** | **9.24 (7.36, 11.61)** | **<0.0001** |
| 8-28 days after | **2.30 (1.75, 3.03)** | **<0.0001** | **2.87 (2.20, 3.74)** | **<0.0001** | **2.12 (1.59, 2.83)** | **<0.0001** |
| 28-56 days after | **2.04 (1.55, 2.67)** | **<0.0001** | **2.35 (1.80, 3.06)** | **<0.0001** | **1.72 (1.29, 2.30)** | **0.0002** |
| **Myocardial infarction** |  |  |  |  |  |  |
| 14-6 days before* | - | - | 3.02 (1.55, 5.89) | 1.00 | - | - |
| 5-1 days before | **4.42 (2.16, 9.05)** | **<0.0001** | **5.37 (2.84, 10.14)** | **<0.0001** | **3.82 (1.81, 8.05)** | **0.0004** |
| 0-7 days after | **4.20 (2.43, 7.25)** | **<0.0001** | **5.36 (3.16, 9.08)** | **<0.0001** | **3.63 (2.04, 6.45)** | **<0.0001** |
| 8-28 days after | 1.22 (0.61, 2.43) | 0.57 | 1.56 (0.80, 3.06) | 0.19 | 1.07 (0.52, 2.19) | 0.85 |
| 28-56 days after | 1.00 (0.49, 2.06) | 1.00 | 1.19 (0.58, 2.43) | 0.63 | 0.86 (0.41, 1.82) | 0.69 |
| **Ischaemic stroke** |  |  |  |  |  |  |
| 14-6 days before* | - | - | 1.73 (0.86, 3.49) | 0.13 |  |  |
| 5-1 days before | 1.80 (0.74, 4.38) | 0.20 | 2.06 (0.92, 4.63) | 0.08 | 1.79 (0.72, 4.45) | 0.21 |
| 0-7 days after | **6.21 (4.22, 9.12)** | **<0.0001** | **7.32 (5.09, 10.53)** | **<0.0001** | **5.81 (3.77, 8.93)** | **<0.0001** |
| 8-28 days after | 0.65 (0.30, 1.40) | 0.27 | 0.76 (0.36, 1.60) | 0.47 | 0.58 (0.26, 1.31) | 0.19 |
| 28-56 days after | 1.06 (0.60, 1.88) | 0.84 | 1.12 (0.64, 1.96) | 0.69 | 0.97 (0.54, 1.77) | 0.93 |
| **Pulmonary embolism** |  |  |  |  |  |  |
| 14-6 days before* | - | - | 2.32 (0.96, 5.65) | 0.06 | - | - |
| 5-1 days before | **7.53 (4.07, 13.94)** | **<0.0001** | **8.08 (4.40, 14.84)** | **<0.0001** | **6.23 (3.28, 11.80)** | **<0.0001** |
| 0-7 days after | **19.88 (14.33, 27.57)** | **<0.0001** | **27.90 (20.79, 37.43)** | **<0.0001** | **16.73 (11.73, 23.86)** | **<0.0001** |
| 8-28 days after | **5.26 (3.55, 7.77)** | **<0.0001** | **7.36 (5.13, 10.56)** | **<0.0001** | **4.20 (2.79, 6.32)** | **<0.0001** |
| 28-56 days after | **4.38 (2.97, 6.46)** | **<0.0001** | **5.66 (3.92, 8.17)** | **<0.0001** | **3.12 (2.07, 4.69)** | **<0.0001** |
| **Deep vein thrombosis** |  |  |  |  |  |  |
| 14-6 days before* | - | - | **3.20 (1.18, 8.69)** | **0.02** | - | - |
| 5-1 days before | **4.32 (1.36, 13.73)** | **0.01** | **3.83 (1.22, 12.07)** | **0.02** | **3.94 (1.20, 12.92)** | **0.02** |
| 0-7 days after | **15.40 (9.29, 25.52)** | **<0.0001** | **17.87 (11.26, 28.35)** | **<0.0001** | **14.51 (8.31, 25.34)** | **<0.0001** |
| 8-28 days after | **3.19 (1.60, 6.33)** | **0.0009** | **3.73 (1.94, 7.17)** | **<0.0001** | **2.59 (1.25, 5.37)** | **0.01** |
| 28-56 days after | 1.77 (0.80, 3.89) | 0.16 | 2.02 (0.93, 4.39) | 0.08 | 1.30 (0.56, 3.01) | 0.53 |

Patients' age quintile was adjusted

IRR: incidence rate ratio

*These pre-test risk intervals were only used for sensitivity analyses on extended pre-test intervals

### **Supplementary Table 2**. E-values for the association between Covid-19 and outcomes

|  | **Non-fatal events** | | **All events** | |
| --- | --- | --- | --- | --- |
|  | **IRR** | **Lower bound of 95% CI** | **IRR** | **Lower bound of 95% CI** |
| **Composite** |  |  |  |  |
| 5-1 days before | 9.01 | 5.85 | 6.88 | 4.44 |
| 0-7 days after | 23.51 | 19.31 | 10.88 | 8.91 |
| 8-28 days after | 5.09 | 3.74 | 2.54 | 1.22 |
| 28-56 days after | 4.03 | 2.94 | 2.45 | 1.71 |
| **Myocardial infarction** |  |  |  |  |
| 5-1 days before | 9.77 | 4.52 | 7.04 | 3.12 |
| 0-7 days after | 9.79 | 5.53 | 3.37 | 1.76 |
| 8-28 days after | - | - | - | - |
| 28-56 days after | - | - | - | - |
| **Ischaemic stroke** |  |  |  |  |
| 5-1 days before | - |  | - |  |
| 0-7 days after | 13.92 | 9.51 | 5.95 | 4.11 |
| 8-28 days after | - | - | - | - |
| 28-56 days after | - | - | - | - |
| **Pulmonary embolism** |  |  |  | - |
| 5-1 days before | 19.39 | 10.31 | 14.42 | 7.73 |
| 0-7 days after | 54.66 | 40.59 | 34.11 | 24.41 |
| 8-28 days after | 14.02 | 9.61 | 6.54 | 4.52 |
| 28-56 days after | - | - | - | - |
| **Deep vein thrombosis** |  |  |  |  |
| 5-1 days before | 8.81 | 2.32 | 7.93 | 2.01 |
| 0-7 days after | 34.37 | 21.49 | 22.51 | 14.08 |
| 8-28 days after | 6.74 | 3.21 | 4.29 | 1.86 |
| 28-56 days after | - | - | - | - |

E-values indicate the minimal association between the time-varying confounder and the outcome that could nullify the association between Covid-19 and outcomes. These were not calculated for non-significant results

### **Supplementary Table 3**. Associations between Covid-19 and non-fatal outcomes by age and deprivation subgroups

|  | **Deprivation** | | | **Age in years** | | |
| --- | --- | --- | --- | --- | --- | --- |
|  | **SIMD ≤2^nd^ quintile** | **SIMD >2^nd^ quintile** |  | **≤65** | **66-80** | **>80** |
|  | **IRR (95% CI)** | **IRR (95% CI)** | **P_interaction_** | **IRR (95% CI)** | **IRR (95% CI)** | **IRR (95% CI)** |
| **Composite** |  |  |  |  |  |  |
| 5-1 days before | 3.26 (2.24, 4.76) | 2.89 (1.91, 4.36) | 0.67 | **6.38 (2.99-13.62)** | **4.20 (2.28-7.71)** | **2.16 (1.02-4.57)** |
| 0-7 days after | 11.04 (8.38, 14.56) | 12.45 (9.49, 16.34) | 0.55 | **24.13 (17.53-33.21)** | **8.15 (5.82-11.40)** | **4.39 (2.97-6.50)** |
| 8-28 days after | 2.23 (1.48, 3.35) | 3.38 (2.38, 4.79) | 0.15 | **7.22 (4.98-10.45)** | 0.93 (0.49-1.76) | 1.10 (0.65-1.85) |
| 28-56 days after | 2.38 (1.66, 3.42) | 2.15 (1.45, 3.19) | 0.72 | **4.45 (2.97-6.66)** | 1.33 (0.81-2.20) | 0.85 (0.49-1.46) |
| **Myocardial infarction** |  |  |  |  |  |  |
| 5-1 days before | 4.38 (2.21, 8.66) | 3.60 (1.88, 6.90) | 0.69 | **11.99 (4.30-33.48)** | **4.29 (1.34-13.74)** | 1.12 (0.16-8.11) |
| 0-7 days after | 6.46 (3.14, 13.30) | 4.48 (2.08, 9.66) | 0.51 | **9.76 (4.15-22.99)** | **4.21 (1.79-9.94)** | 1.75 (0.54-5.60) |
| 8-28 days after | 1.92 (0.78, 4.75) | 1.26 (0.46, 3.44) | 0.57 | **3.65 (1.44-9.28)** | 0.38 (0.05-2.79) | 0.87 (0.27-2.77) |
| 28-56 days after | 1.31 (0.48, 3.59) | 1.09 (0.40, 2.98) | 0.80 | 1.20 (0.29-4.98) | 0.65 (0.16-2.67) | 0.96 (0.34-2.67) |
| **Ischaemic stroke** |  |  |  |  |  |  |
| 5-1 days before | 2.47 (1.31, 4.68) | 1.14 (0.42, 3.09) | 0.22 | - | 1.76 (0.43-7.18) | 1.93 (0.62-6.06) |
| 0-7 days after | 6.50 (3.87, 10.91) | 8.16 (4.92, 13.54) | 0.54 | **15.03 (7.19-31.42)** | **5.65 (3.10-10.29)** | **3.56 (1.92-6.59)** |
| 8-28 days after | 0.76 (0.28, 2.07) | 0.68 (0.22, 2.14) | 0.89 | 0.73 (0.10-5.41) | 0.20 (0.03-1.48) | 0.74 (0.30-1.82) |
| 28-56 days after | 0.61 (0.22, 1.64) | 1.64 (0.83, 3.25) | 0.12 | 1.62 (0.49-5.37) | 0.66 (0.24-1.83) | 0.60 (0.24-1.47) |
| **Pulmonary embolism** |  |  |  |  |  |  |
| 5-1 days before | 4.72 (2.39, 9.32) | 4.40 (2.04, 9.50) | 0.89 | **10.00 (3.12-32.08)** | **11.21 (4.84-25.96)** | 3.76 (0.91-15.58) |
| 0-7 days after | 25.44 (16.80, 38.54) | 31.31 (20.59, 47.61) | 0.49 | **52.33 (33.5-81.74)** | **16.42 (9.70-27.80)** | **7.56 (3.50-16.36)** |
| 8-28 days after | **4.32 (2.32, 8.07)** | **11.23 (7.14, 17.67)** | **0.02** | **14.63 (8.92-24.00)** | **2.98 (1.40-6.34)** | 1.75 (0.62-4.93) |
| 28-56 days after | 6.66 (4.17,10.65) | 4.47 (2.46, 8.12) | 0.32 | **7.80 (4.54-13.38)** | **2.90 (1.46-5.74)** | 2.16 (0.90-5.18) |
| **Deep vein thrombosis** |  |  |  |  |  |  |
| 5-1 days before | 1.79 (0.44, 7.36) | 5.51 (2.20, 13.83) | 0.19 | - | 3.15 (0.42-23.32) | **6.36 (1.49-27.14)** |
| 0-7 days after | 15.80 (8.23, 30.33) | 20.94 (10.85, 40.38) | 0.55 | **25.39 (12.03-53.56)** | **10.74 (4.56-25.29)** | **8.26 (3.28-20.78)** |
| 8-28 days after | 4.27 (1.83, 9.97) | 3.39 (1.21, 9.44) | 0.74 | **7.91 (3.42-18.26)** | - | 1.85 (0.53-6.46) |
| 28-56 days after | 2.30 (0.83, 6.41) | 2.11 (0.65, 6.79) | 0.92 | **3.39 (1.18-9.76)** | 1.34 (0.32-5.73) | 0.52 (0.07-3.90) |

Patients' age quintile was adjusted

IRR: incidence rate ratio

### **Supplementary Table 4**. Associations between Covid-19 and outcomes in cohort analysis

|  | **N** | **Event** | **HR (95% CI)** | **P** |
| --- | --- | --- | --- | --- |
| Composite | 211895 | 2785 | 7.81 ( 7.18, 8.49) | <0.0001 |
| MI | 224991 | 439 | 3.31 ( 2.59, 4.22) | <0.0001 |
| Ischaemic stroke | 224799 | 387 | 4.40 ( 3.44, 5.63) | <0.0001 |
| Pulmonary embolism | 225598 | 259 | 24.07 (18.49, 31.33) | <0.0001 |
| Deep vein thrombosis | 225873 | 107 | 10.45 ( 7.02, 15.56) | <0.0001 |

HR: hazard ratios estimated in Cox regression analysis after adjusting for age, sex, and deprivation
